# Efficacy and safety of sustained release ashwagandha root extract capsules on muscle strength and cardiorespiratory endurance in recreationally active individuals: A randomized, double-blind, placebo-controlled trial

**DOI:** 10.64898/2026.06.29.26356803

**Authors:** Shefali Thanawala, Rajat Shah, Alphy Lopes, Girish Rudrappa, Prabakaran Desomayanandanam, Sahitya Srinivas

**Affiliations:** Medical Science & Research, Nutriventia Private Limited, Mumbai, Maharashtra, 400069 India; Executive Management, Nutriventia Private Limited, Mumbai, Maharashtra, 400069 India; Orthopedics Department, Rajalakshmi Hospital and Research Center, Bangalore, Karnataka, 560097 India; Clinical Operations, Invitro Research Solutions Private Limited, Bangalore, Karnataka, 560092 India

**Keywords:** cardiorespiratory endurance, muscle damage, recovery, muscle strength, physical performance, quality of life, Withania somnifera

## Abstract

**Introduction:** Previous studies suggest that conventional Ashwagandha (*Withania somnifera*) root extracts may improve physical performance in both athletic and non-athletic populations. However, evidence on sustained-release ashwagandha formulations is lacking. This randomized double-blind, placebo-controlled trial evaluated the safety and efficacy of sustained-release ashwagandha root extract (AshwaSR) 300 mg in improving muscle strength in recreationally active healthy individuals.

**Methods:** Eligible participants aged 25-50 years received AshwaSR 300 mg or placebo capsules once daily for 90 days. Changes in one-repetition maximum (1-RM) bench press, maximal oxygen uptake (VO_2_max), serum testosterone, creatine kinase (CK) and lactate dehydrogenase (LDH) concentrations, and quality of life (QoL using SF-12 questionnaire) from baseline till study end were evaluated.

**Results:** Of 84 enrolled participants, 83 completed the study (AshwaSR, n=41; Placebo, n=42). Compared with placebo, participants receiving AshwaSR demonstrated significantly greater improvements in muscle strength and cardiorespiratory endurance starting from day 30 (1-RM: 4.69 vs 2.23 kg; VO_2_max: 9.99 vs 6.64 mL/kg/min; *p*<0.001), and sustained till day 91 (1-RM: 15.31 vs 7.33 kg; VO_2_max: 30.56 vs 18.14 mL/kg/min; *p*<0.001). In the AshwaSR group, total testosterone levels significantly increased from baseline to day 90 among men (mean increase = 121.43 ng/dL; *p*=0.024); while the improvement in testosterone levels among women was non-significant (13.34 ng/dL; *p*=0.545) and remained within normal reference ranges. The favorable numerical changes in CK and LDH concentrations were observed in AshwaSR group at the end of the study. AshwaSR group reported significantly improved quality of life at day 91 compared to placebo group (Mental component: 7.72 vs −4.66; *p*<0.001; Physical component: 5.23 vs −1.91; *p*<0.001). Reported adverse events in both groups were mild and transient in nature.

**Conclusion:** Three-months supplementation with AshwaSR 300 mg/day was safe and effective in improving muscular strength, cardiorespiratory endurance, and overall wellbeing in recreationally active healthy adults.

**Clinical Trial Registry of India (CTRI) registration number:** CTRI/2025/04/084383

## 1 Introduction

As the pursuit of longevity and optimal health gains momentum, physical fitness and independence have become the fundamental pillars of lifelong well-being. Skeletal muscle strength is a key determinant of healthy ageing, physical performance, functional independence, metabolic health, and long-term quality of life. Evidence suggests that both muscle strength and the processes involved in its development are closely linked to healthy ageing (1, 2). Cardiorespiratory fitness is another critical component of physical health, with maximal oxygen uptake (VO₂max) reflecting the integrated capacity of the cardiovascular and pulmonary systems to deliver, and muscular systems to utilize oxygen during exercise. Higher VO₂max is strongly associated with improved endurance performance and favorable cardiometabolic outcomes. Thus, muscle strength and cardiorespiratory fitness together play a central role in the prevention and delaying of the progression of chronic diseases, underscoring their value as key indicators of overall health status (3–6).

Physical exercise, whether in the form of resistance training, aerobic training or a combination of both, is widely recognized for its ability to enhance muscular strength and cardiorespiratory endurance. In addition to enhancing physical performance, regular exercise contributes to improved body composition and lowers the risk of chronic non-communicable diseases (7). However, structured strength training is often associated with challenges such as muscle soreness, fatigue, and delayed recovery, instrumental in poor adherence and resultant failure to achieve the targeted health goals. Particularly, repeated or strenuous exercise can induce transient muscle damage characterized by microtears of sarcomeres, inflammation, and oxidative stress. Such exercise-induced muscle damage is commonly reflected by elevations in circulating biomarkers such as creatine kinase (CK) and lactate dehydrogenase (LDH), which are indirect indicators of muscle damage and recovery status (8–11). However, as this entire process of muscle damage followed by recovery is rather a physiological process, nutraceuticals and dietary supplements that can help support recovery while improving training adaptations have become an area of growing interest in sports nutrition and for athletes and fitness enthusiasts. Special attention is focused towards botanicals, primarily due to the growing consumer preference in evidence-based natural approaches for achieving optimum performance targets (12).

Ashwagandha (*Withania somnifera* (L.) Dunal), a well-recognized herb used in traditional Ayurvedic practice, has attracted substantial scientific interest owing to its adaptogenic, anti-stress, anti-inflammatory, antioxidant, and immunomodulatory properties. Standardized root extracts of ashwagandha contain bioactive phytoconstituents, primarily steroidal lactones, known as withanolides, which have been investigated for their effects on stress physiology, neuromuscular performance, and endocrine balance. Over the past decade, evidence from several clinical studies has highlighted the potential role of ashwagandha extract as an ergogenic aid, with supplementation shown to improve muscle strength and recovery outcomes in healthy adults and athletes (13–15). Emerging evidence also supports the role for ashwagandha in cardiorespiratory performance. A systematic review and meta-analysis reported that ashwagandha supplementation significantly improved VO₂max in healthy adults and athletes, although the authors noted heterogeneity and the need for additional well-designed trials. These trials have evaluated conventional immediate-release preparations administered once or twice daily in the doses ranging from 500 mg – 1250 mg (16). However, high dosing frequency can affect long-term compliance, which can, in turn, affect the results (17, 18).

Ashwagandha root-only extract sustained-release (SR) capsules 300 mg (AshwaSR - Ashwanova™, formerly known as Prolanza™) were developed to address this issue and to improve dosing convenience and enhance compliance. The sustained-release delivery technology used in this formulation allows the gradual release of the actives over an extended period thereby providing prolonged therapeutic effect with a single daily dose. The reduced dosing frequency helps improve user convenience and long-term compliance compared with conventional immediate-release formulations. The multidimensional clinical potential of AshwaSR has been supported by a systematic stepwise approach of translational research wherein its antioxidant and anti-inflammatory effects were demonstrated in both *in vitro* and *in vivo* studies. Also, the sustained-release profile of the formulation was confirmed in the *in vivo* pharmacokinetic study. The efficacy of the formulation was further evaluated in a chronic unpredictable stress animal model (19, 20). These preclinical pharmacodynamic and pharmacokinetic findings were subsequently corroborated through clinical trials in healthy human participants. In a randomized clinical trial, supplementation with AshwaSR 300 mg capsules for 3 months significantly improved cognitive performance, psychological well-being, sleep quality, and stress parameters in healthy adults (21). Furthermore, in a randomized controlled trial involving healthy stressed adults, supplementation with AshwaSR for 60 days at low (150 mg) as well as standard dose (300 mg) was effective in reducing perceived stress and improving sleep quality, eating behavior, and psychological well-being (22).

These findings from clinical trials, along with established evidence on potential benefits of Ashwagandha extracts in sports nutrition, provide a rationale for exploring the efficacy of AshwaSR in physical fitness outcomes. Although several clinical trials have investigated the effects of conventional ashwagandha extracts on muscle strength and aerobic performance, clinical evidence regarding sustained-release ashwagandha formulations is scarce, representing a significant gap in the current literature. Therefore, the present randomized controlled trial was conducted to primarily evaluate the safety and efficacy of AshwaSR 300 mg, administered for a period of 90 days, on muscle strength in recreationally active healthy individuals. Additionally, we assessed its effects on cardiorespiratory endurance, muscle recovery biomarkers, testosterone concentrations, and quality of life (QoL). Through assessment of all these outcomes, this study sought to provide a comprehensive evaluation of sustained-release ashwagandha root extract as a supportive intervention for sports nutrition and exercise adaptation.

## 2 Materials and methods

### 2.1 Study design and ethical considerations

This randomized, double-blind, placebo-controlled, prospective, single-center, parallel group clinical trial was conducted at Rajalakshmi Hospital and Research Center, Bangalore, Karnataka, India between April 2025 and November 2025. The trial protocol followed the ethical guidelines set forth by the Declaration of Helsinki for research involving human participants. The trial was conducted in compliance with Good Clinical Practice (GCP), the Helsinki Declaration Standards, the New Drug and Clinical Trial (NDCT) Rules 2019, and Indian Council of Medical Research (ICMR) codes. Both trial protocol and informed consent form were approved by Rajalakshmi Hospital Institutional Ethics Committee, Bengaluru (Bangalore), India (Re-registration number: ECR/809/Inst/KA/2017/RR-25; Approval date: 01.09.2025). This trial was registered with the Clinical Trials Registry-India (CTRI) on 08 April 2025 (CTRI number: CTRI/2025/04/084383). All participants provided written informed consent prior to initiation of the trial.

### 2.2 Study Population eligibility

Study participants were recruited through a site-based screening approach. Eligibility was determined according to predefined inclusion and exclusion criteria to ensure a homogeneous population of recreationally active individuals. Eligible participants included men and non-pregnant women aged between 25 and 50 years, with a body mass index (BMI) between 25.0 and <29.9 kg/m², who were recreationally active, gym going, and at the initial stages of strength training (<1 month).

Additional inclusion criteria were i) healthy participants determined based on a medical history, physical examination, and the clinical judgment of the principal investigator (PI), who had clinical laboratory parameters at screening within reference ranges or deemed clinically insignificant by the PI; ii) participants who agreed to avoid the intake of any dietary or ergogenic supplements, including protein formulations, metabolic enhancers, or anabolic steroids that could influence muscular adaptations and who were ready to maintain their habitual diet without significant modifications throughout the study duration. Women of reproductive potential were required to adopt a medically acceptable form of contraception throughout the study duration. Women who were amenorrheic for at least one year or had undergone hysterectomy and/or bilateral oophorectomy were considered non-childbearing. Participants were required to adhere to a standardized exercise regimen consisting of five training days per week within a seven-day cycle, incorporating alternating resistance and aerobic training sessions followed by two rest days. To minimize confounding influences, participants were required to be free from dependence on alcohol, tobacco, or other substances of abuse and to abstain from their use for the duration of the study. Lastly, adherence to study procedures and protocol requirements was mandatory for all eligible participants.

The exclusion criteria were participants i) with hypersensitivity to any of the ingredients of the study products; having any significant uncontrolled systemic illness (including but not limited to uncontrolled hypertension, chronic liver or renal disease, poorly controlled diabetes mellitus, congestive heart failure, coronary artery disease, musculoskeletal disorders etc.); ii) who were concurrently using supplements for stress/anxiety/sleep/any other indications, herbal or pharmaceutical preparations; iii) who were on any medication/treatment for infertility/enhancement of sexual function within 6 months prior to study enrolment. Additionally, participants who had experienced a body weight reduction exceeding 5 kg within three months prior to screening, or those with recent (within six months) orthopedic injury or surgical intervention were excluded. Participation in other clinical trials involving investigational or marketed products within three months prior to enrollment or those with any condition that, in the opinion of the PI, could interfere with study participation were also excluded from the trial.

### 2.3 Treatment allocation, Randomization and Blinding

Eligible participants were randomized to receive either AshwaSR 300 mg capsules or placebo capsules in accordance with a pre-established randomization schedule. Test product, AshwaSR (Ashwanova™, formerly known as Prolanza™, Nutriventia Private Limited, Mumbai, Maharashtra, India) 300 mg capsules contained sustained-release *Withania somnifera* root extract (standardized to contain not less than 4% of total USP withanolides) and permitted excipients. Placebo capsules contained only inactive substances. To maintain treatment concealment, placebo and test product capsules were identical in appearance, including color, size, and shape. The investigational products (IPs) were manufactured by Nutriventia Private Limited.

Randomization was implemented using a computer-generated sequence prepared by an independent study statistician using Statistical Analysis System (SAS^®^) software (version 9.4). Treatment assignments were linked to unique randomization numbers, which were allocated sequentially. An independent unblinded pharmacist maintained the allocation sequence and ensured blinding of study products by removing original product labels and replacing them with blinded labels corresponding to the assigned randomization codes. Access to the randomization codes was not disclosed to the investigators or study personnel until study completion. All IP containers were assigned sequential identification numbers and labelled with a common batch number across treatment groups. Upon confirmation of participant eligibility, study personnel contacted the unblinded pharmacist to obtain the corresponding randomization number and assigned container for dispensing. This approach ensured that both participants and investigators remained blinded to treatment allocation throughout the study. After study completion, randomization codes were shared with the data management team for final statistical analysis.

### 2.4 Study procedure

The study was conducted for approximately 107 days which included screening period of 16 days (Visit 1 [day −16 to −1]), intervention period of 90 days (Visit 2 [base-line/day 01], Visit 3 [day 02], Visit 4 [day 03], Visit 5 [day 30 ± 2], Visit 6 [day 60 ± 2], Visit 7 [day 89 ± 2], Visit 8 [day 90]) and Visit 9 (day 91) one day after completion of study intervention for post-study assessments (Figure 1). The screening period included ac-climatization period of 14 days where the participants performed pre-defined workout regimen.

**Figure 1:**
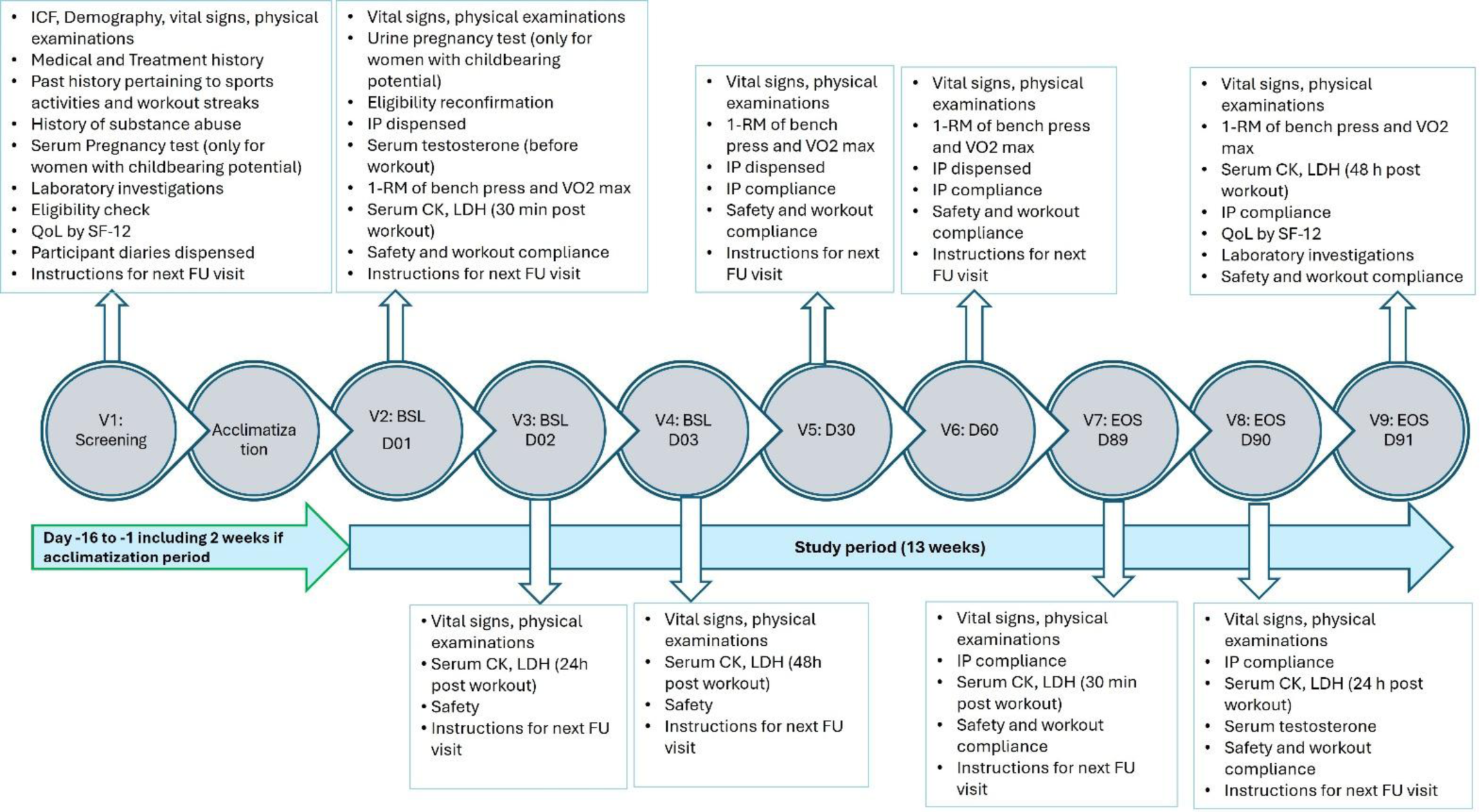
Study flowchart. Abbreviations: CK, creatine kinase; EOS, end of study; FU, follow-up; IP, investigational product; LDH, lactate dehydrogenase; QoL, quality of life; 1-RM, one-repetition maximum; SF-12, 12-Item Short Form Health Survey; SR, sustained-release; VO_2_max, maximal oxygen uptake.

A series of screening evaluations performed after taking informed consent included collection of participant data pertaining to demography including age, gender, body weight, height, BMI, medical and treatment history details. Additionally, participants underwent a general physical examination, vital signs assessments (body temperature, pulse rate, respiratory rate and blood pressure), and laboratory assessments (including hematology, liver function tests, renal function tests, and urine analysis). On baseline visit (day 01), reconfirmation of eligibility criteria, along with physical examination, vital signs assessments, urine pregnancy test in women of childbearing potential to rule out pregnancy were conducted. At baseline visit, all eligible participants were assessed for muscle strength using one-repetition maximum (1-RM) of bench press, cardiorespiratory endurance using VO_2_max parameter and then proceeded to their respective daily workout training.

Participants’ blood samples were collected prior to the workout for measurement of baseline serum testosterone concentrations and 30 minutes post-workout for measurement of serum CK and LDH concentrations. Participants were randomized to receive either AshwaSR 300 mg capsules or placebo capsules. The IPs were dispensed to participants in a bottle containing 32 capsules at the baseline/randomization visit (day 01), and subsequent interim visits (day 30 and day 60). They were advised to consume one capsule daily, orally with water, following breakfast in the morning for 90 days. On subsequent interim visits on day 02 (24 h post day 01 workout) and 03 (48 h post day 01 workout), blood samples were collected to measure serum CK and LDH concentrations. Quality of life was assessed on screening visit and at the end of study (Visit 9) using 12-Item Short Form Health Survey (SF-12) questionnaire. Muscle strength and cardiorespiratory endurance assessments were repeated at interim visits (day 30, day 60) and end of the study (day 91). Similar to baseline visits, at the end of study period, serum CK and LDH concentrations were evaluated at day 89 (30 minutes after workout), day 90 (24 h post day 89 workout), and day 91 (48 h post day 89 workout). Serum testosterone measurements were repeated on day 90.

Participant diary was given to every participant at screening visit, day 30, and day 60 to record intake of IPs, daily work-out regimen details, adverse events (AEs) experienced, concomitant medications taken, if any. They were instructed to fill these diaries regularly on daily basis. These diaries were reviewed by the study team at each visit for workout and IP compliance, along with safety assessment. Unused IPs were collected from participants on days 30, 60, and 90. Treatment compliance was considered adequate if participants consumed ≥80% of the scheduled IP doses.

Assessment of physical ex-aminations, vital signs, concomitant medication use and AE monitoring were conducted at all study visits while laboratory clinical assessments were conducted at screening and end of the study (day 91). Treatment-emergent adverse events (TEAEs) were recorded and evaluated as a part of safety assessment. Participants were instructed to promptly inform the PI or authorized study personnel if any illness occurred that required the use of additional medications during the study period. The subject’s and physician’s global assessment of therapy was assessed at the end of the treatment (day 90) to evaluate the general acceptability and value of in-tended benefits from the study interventions.

#### 2.4.1 Workout regimen details

A structured and progressive resistance training program was implemented to standardize physical activity across participants and to train the targeted muscle groups of both the upper and lower body. The exercise regimen was designed in alignment with established recommendations from the National Strength and Conditioning Association, incorporating multiple exercises predefined to be performed with appropriate resistance, organized into sets and repetitions to promote muscular adaptation. All participants were required to train for a two-week acclimatization phase (which was completed within the 16 days of screening period) followed by the 13-weeks of intervention period. A uniform 2-week workout regimen was implemented during the acclimatization phase to familiarize the participants with study workout regimen. During this phase, exercise volume and intensity were gradually introduced to ensure participant familiarization with movement patterns and equipment. Participants who completed acclimatization workout training were further randomized in the study. One workout regimen week consisted of five continuous workout days alternating between resistance training (three days) and aerobic training (two days) ending with two days’ rest. Detailed workout regimen is summarized in table 1.

**Table 1:**
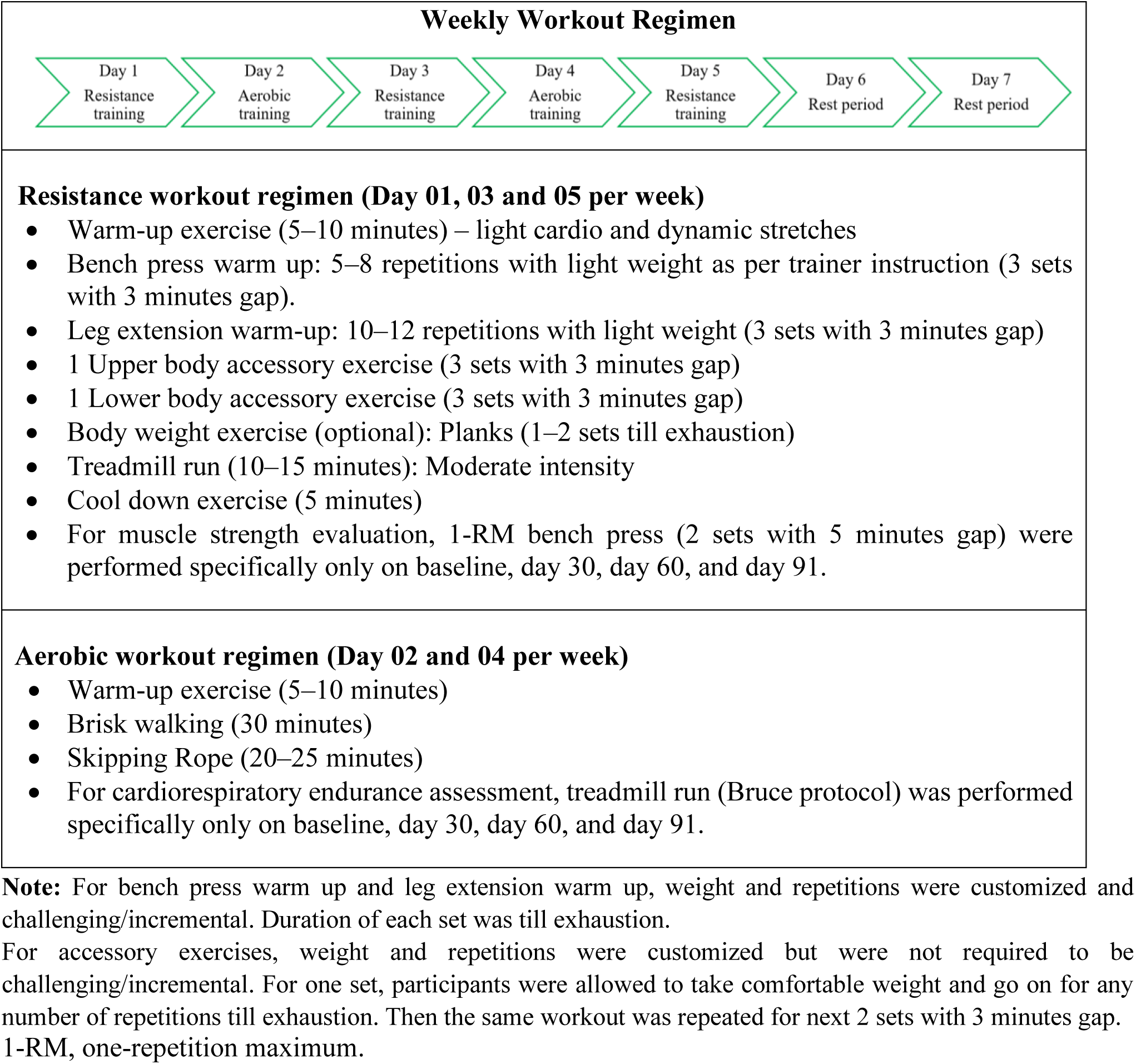
Weekly workout regimen.

### 2.5 Endpoints

The primary efficacy endpoint was to evaluate and compare the mean change in muscle strength measured by 1-RM of bench press load from baseline to end of the study (day 91), between the AshwaSR and placebo groups. The secondary endpoints were to evaluate and compare change in muscle strength at interim follow-up visits (at days 30 and 60) from baseline between study groups, change in VO_2_max from baseline to day 30, 60, and 91, change in serum testosterone concentrations from baseline to end of treatment (day 90), change in muscle damage markers including serum CK and LDH concentrations from baseline (day 1, 2, and 3) to the end of the study (day 89, 90, and 91). Additionally, we evaluated and compared change from baseline to end of the study (day 91) in QoL using SF-12 questionnaire; comparison of Subject’s and Physician’s global assessment of therapy at day 90 between the groups was conducted. Safety assessments were conducted which included evaluation of incidence, frequency, and severity of AEs and TEAEs throughout the study duration.

### 2.6 Study assessments

#### 2.6.1 Muscle strength – 1-RM bench press load

Muscle strength was assessed using the 1-RM, defined as the maximum weight an individual can lift for a single repetition of a given exercise (23). 1-RM is a widely accepted and validated measure of maximal muscle strength, as recognized by the American College of Sports Medicine, and is commonly employed in healthy populations (24). In the present trial, upper body muscle strength was evaluated using the bench press exercise, which is considered a reliable and time-efficient method for assessing maximal strength in physically active individuals compared to lower-body alternatives such as leg extension. 1-RM was estimated using a submaximal approach based on Epley’s formula (23). Participants were instructed to lift a challenging but safe weight for multiple repetitions until volitional exhaustion, and the 1-RM was calculated using the following equation:

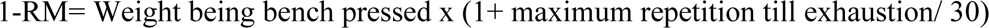

Assessments were conducted at baseline (day 01), day 30, day 60, and at the end of the study (day 91). Higher 1-RM values were interpreted as greater muscle strength. The assessment procedure was standardized across study visits. All assessments were conducted under supervision to ensure proper lifting techniques, minimize risk of injury, and maintain consistency across participants.

#### 2.6.2 Cardiorespiratory endurance (VO_2_max)

Cardiorespiratory endurance was assessed by estimating VO_2_max, which represents the maximum capacity of an individual to uptake, transport, and utilize oxygen during intense exercise. It is a key indicator of aerobic and cardiovascular fitness. Higher VO_2_max values reflect superior cardiorespiratory endurance and aerobic capacity. VO_2_max was evaluated using the Bruce treadmill protocol, a validated indirect method for estimating aerobic capacity (25–27). The test was conducted on a motorized treadmill using an incremental design consisting of successive stages, each lasting 3 minutes to allow attainment of steady-state heart rate. With each stage, both the speed and incline (grade) of the treadmill were progressively increased until volitional exhaustion, thereby imposing greater cardiovascular demand. Treadmill-based assessment was selected due to its ability to elicit higher exercise intensity and greater cardiovascular stress compared to other modalities, thereby providing a robust estimate of aerobic performance (28). To ensure accuracy of VO_2_max estimation, heart rate stabilization within each stage was monitored. VO_2_max was evaluated at baseline (day 01), day 30, day 60, and day 91.

#### 2.6.3 Quality of life (SF-12)

Health-related QoL was evaluated using the SF-12, a standardized, validated and self-reported questionnaire. It consists of 12 items from eight health domains, including physical functioning, physical role, bodily pain, general health, vitality, social functioning, emotional role, and mental health. The questionnaire generates two composite scores: the Physical Component Summary (PCS) and the Mental Component Summary (MCS), which reflect overall physical and mental health status, respectively. It is widely used in clinical trials due to its psychometric validity in both general and patient populations (29). The SF-12 assessment was completed by the participants at baseline and on day 91. A higher score reflects better health and quality of life.

#### 2.6.4 Serum biomarkers (CK, LDH and testosterone)

Serum CK and LDH were assessed as markers of exercise-induced muscle damage and recovery. Blood samples were collected at 30 minutes, 24 hours, and 48 hours post-exercise at baseline and at the end of the study. Changes in CK and LDH levels were evaluated and compared between the study groups to determine the effect of the intervention on muscle damage and subsequent recovery, as well as the potential adaptation to resistance training. Additionally, serum testosterone levels were measured at baseline and at the end of the treatment (day 90) as increased physical performance and resistance training are often associated with modulation of testosterone levels, which may influence muscle strength and endurance. The laboratory panel for serum testosterone assessment included measurements of free testosterone and total testosterone levels.

### 2.7 Sample size calculation

The Sample size was determined based on the previously published evidence reporting improvements in muscle strength assessed by 1-RM bench press (13). We assumed that the test product (AshwaSR) in the present study will result in a minimum difference of 10 kg from baseline relative to placebo. Assuming a standard deviation of 16.72 kg, a total of 76 evaluable participants (38 participants per treatment) were required to identify the difference with 80% power and at a two-sided significance level of 0.05. Considering 10% dropout rate and non-compliance rate, a total of 84 participants (equally randomized as 42 participants per treatment) were required to be enrolled in the study.

### 2.8 Statistical analysis

A compliance to study interventions of ≥ 80% was considered acceptable and was considered for efficacy evaluation and statistical analysis. The statistical analysis was conducted using the SAS^®^, Version 9.4. Baseline characteristics were summarized using mean and standard deviation (SD) for continuous variables and using number and percentages for categorical variables.

The continuous variables of the study outcome measures were compared between the study groups for the baseline adjusted changes by a mixed effect model ANCOVA with baseline observation as covariate. The mixed effect model also accounted for the adjustment of homogeneity of variances. The statistical significance of relative changes at post-baseline assessment timepoints from baseline of each outcome measure was evaluated by paired t-test. In addition, Student t-test for independent variables was used as applicable to evaluate the differences between study groups. All analyses were done at a 0.05 significance level wherein *p* < 0.05 was considered statistically significant.

## 3 Results

Of the 87 participants screened, 84 eligible participants were enrolled (Screen failure, n = 03) and randomized to either AshwaSR (n = 42) or placebo (n = 42) groups. One participant was lost to follow-up, and a total of 83 participants including 54 men and 30 women (AshwaSR, n = 41 and Placebo, n = 42) completed the study (Figure 2). Demographic characteristics of the study participants between the study groups were comparable at baseline and are summarized in table 2.

**Figure 2:**
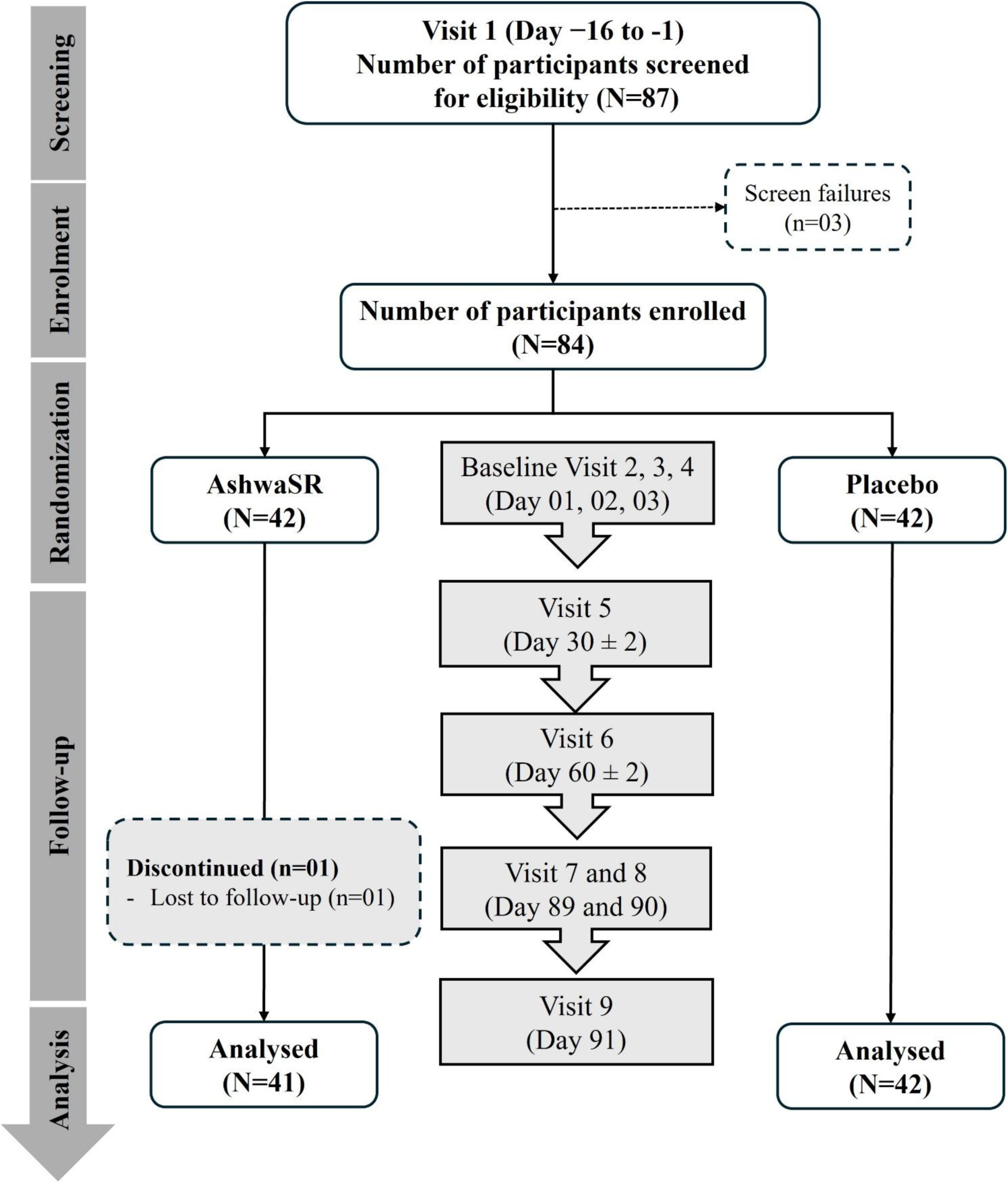
CONSORT flow diagram.

**Table 2:**
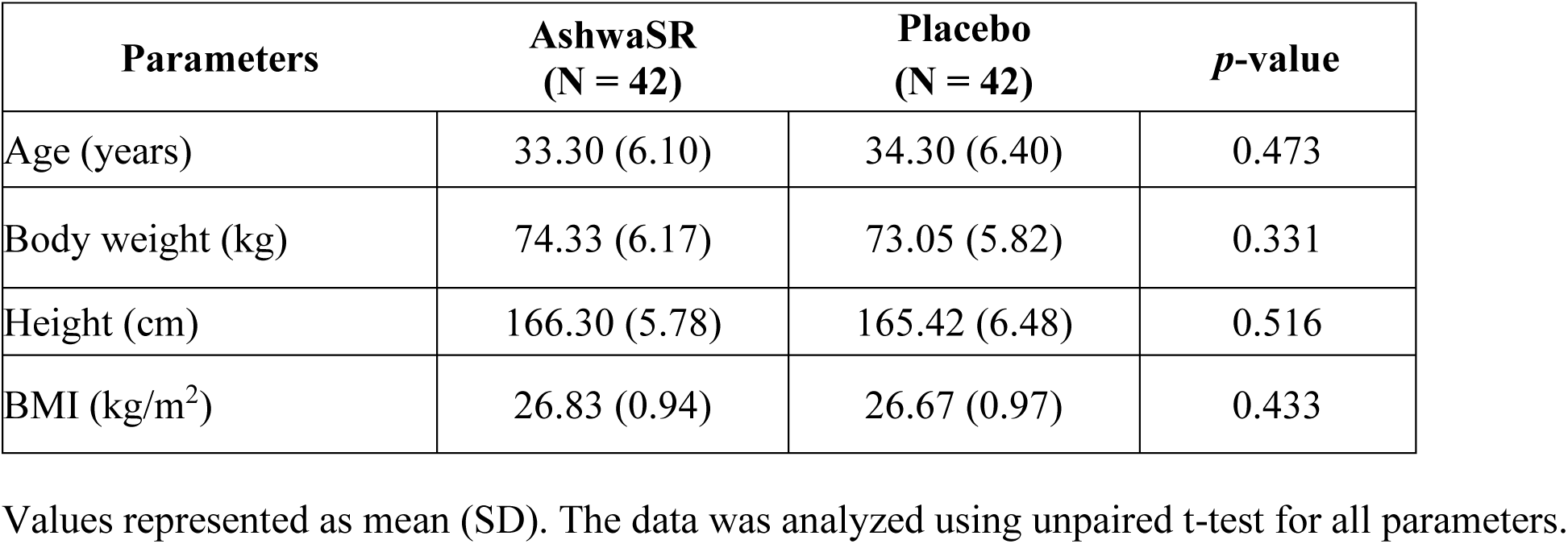
Demographic characteristics at baseline.

### 3.1 Efficacy outcomes

3.1.1 **Muscle strength by 1-RM of bench-press**

Participants from both groups reported statistically significant increase in mean muscle strength from day 30 onwards until day 91, compared to baseline (*p* < 0.001). A between group comparison demonstrated that mean improvement in 1-RM bench press from baseline to day 30, 60, and 91 in AshwaSR group were statistically significantly higher compared to that observed in placebo group (*p* < 0.001) (Figure 3). The magnitude of increase in muscle strength from baseline at day 30, 60, and 91 was 79.75%, 163.67%, and 260.40% respectively, in AshwaSR group and 36.36%, 74.45%, and 119.31%, respectively, in placebo group.

**Figure 3:**
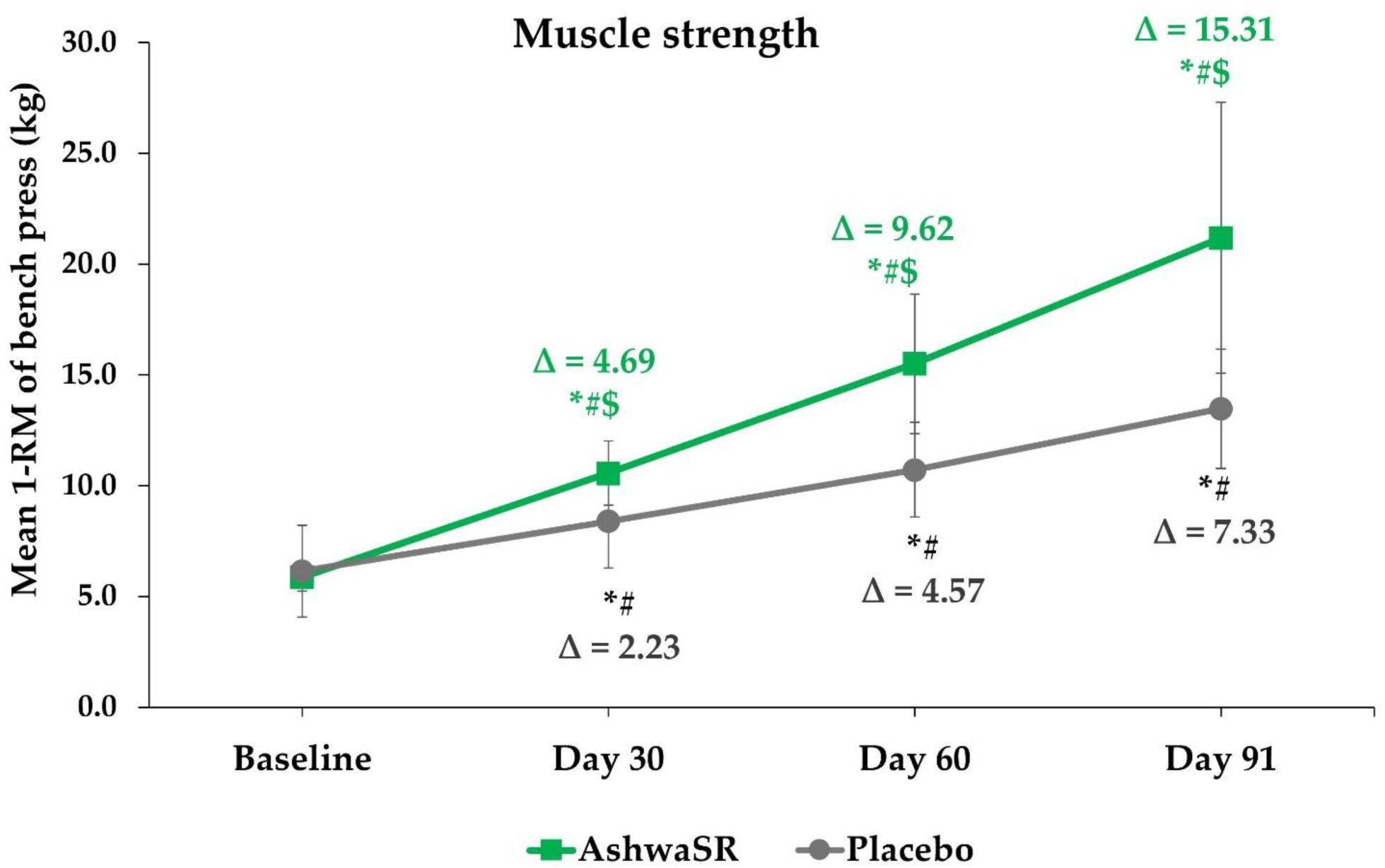
Comparison of mean change in muscle strength from baseline to end of the study (day 91) between AshwaSR and placebo. Data presented as mean and SD (Error bars represent SD). Δ represents mean change from baseline to the follow-up timepoints (days 30, 60, and 91). * represents *p*-value < 0.05 which indicates statistically significant difference. *^#^ shows statistically significant difference in mean scores between baseline and each follow-up timepoint (*p*-value derived from Paired t-test). *^$^ shows statistically significant difference in the mean change (from baseline to all follow-up timepoints) between AshwaSR and placebo (*p*-value derived from ANCOVA). Abbreviations: 1-RM, repetition max; SD, standard deviation.

#### 3.1.2 Cardiorespiratory endurance by VO_2_max

The mean (SD) VO_2_max statistically significantly increased from baseline to day 30, 60, and 91 in both study groups. A comparison of mean change in VO_2_max values between groups showed that improvements from baseline observed in VO_2_max at days 30, 60, and 91 were statistically significantly higher in participants administered with AshwaSR, compared to those observed in participants from placebo group (*p* < 0.001). The magnitude of increase in VO_2_max from baseline at day 30, 60, and the end of the study (day 91) was 47.44%, 85.70%, and 145.12%, respectively, in AshwaSR group and 32.40%, 54.03%, and 88.54% in placebo group (Figure 4).

**Figure 4:**
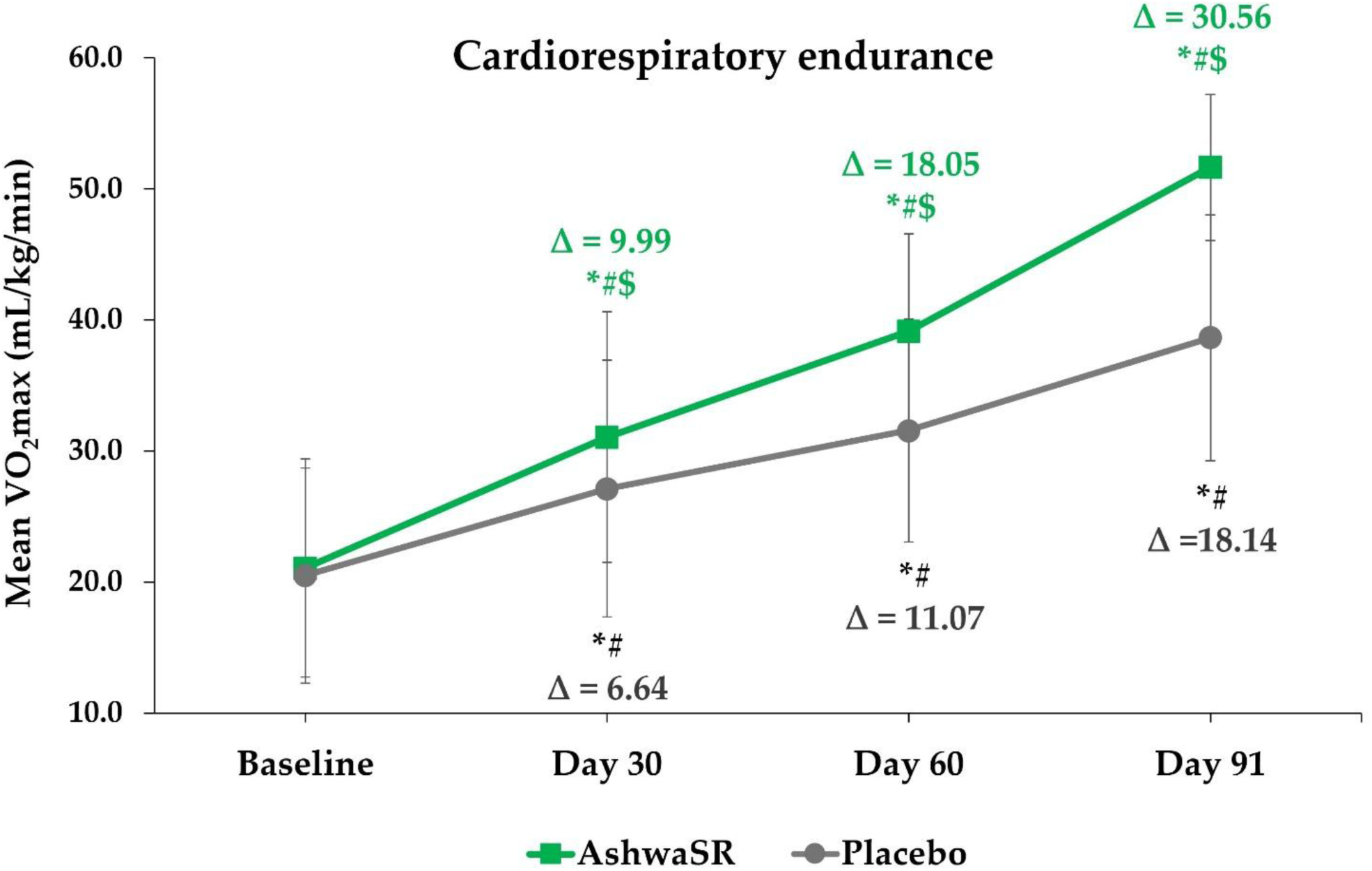
Comparison of mean change in cardiorespiratory endurance (VO_2_max) from baseline to end of the study (day 91) between AshwaSR and placebo. Data presented as mean and SD (Error bars represent SD). Δ represents mean change from baseline to the follow-up timepoints (days 30, 60, and 91). * represents *p*-value < 0.05 which indicates statistically significant difference. *^#^ shows statistically significant difference in mean scores between baseline and each follow-up timepoint (*p*-value derived from Paired t-test). *^$^ shows statistically significant difference in the mean change (from baseline to all follow-up timepoints) between AshwaSR and placebo (*p*-value derived from ANCOVA). Abbreviations: VO_2_max, maximum oxygen capacity; SD, standard deviation.

#### 3.1.3 Serum testosterone

For the analysis of serum testosterone levels, a sex-stratified approach was used due to natural differences in clinical reference ranges between men and women.

In male participants, at day 90, the AshwaSR group demonstrated a statistically significant increase from baseline in total testosterone levels (mean change = 121.43 ng/dL, *p* = 0.024), however, no significant improvement in total testosterone was observed in the placebo group (mean change = 68.43 ng/dL; *p* > 0.05). In both study groups, free testosterone concentrations were statistically significantly improved with mean change of 2.99 ng/dL (*p* = 0.006) in AshwaSR group and 2.32 ng/dL (*p* = 0.007) in placebo groups. The percentage increase from baseline to day 90 in the AshwaSR group was 38.47% and 51.01% for total and free testosterone, respectively, compared to 20.61% and 39.91% in the placebo group. However, between group differences were not statistically significant for both total and free testosterone concentrations (*p* > 0.05).

In female participants, within group analysis showed that at day 90, the mean concentrations of total and free testosterone were higher in the AshwaSR group, com-pared to baseline (*p* > 0.05). In contrast, the placebo group demonstrated a statistically significantly higher concentration of total (*p* = 0.003) and free (*p* = 0.002) testosterone at day 90 than baseline concentrations. The mean increase in total testosterone concentration was significantly higher in placebo group (28.61 ng/dL) than that observed in AshwaSR group (13.34 ng/dL) (*p* = 0.021). The mean increase from baseline in free testosterone was 0.27 ng/dL in AshwaSR group and 0.47 ng/dL in the placebo group (*p* > 0.05) The percentage increase from baseline to day 90 in the AshwaSR group was 44.43%, and 59.68% for total and free testosterone, respectively, compared to 73.25%, and 76.26% in the placebo group.

#### 3.1.4 Muscle damage biomarkers

Evaluation of muscle damage markers showed that at the end of the study, a reduction in the mean change of CK concentrations was observed from 0 h to 48 h in the AshwaSR group (Mean difference at 0 h =16.69 U/L, 24 h =11.81 U/L, and 48 h = −48.24 U/L). In contrast, in the placebo group the mean difference in CK concentrations was plateaued at 0 h and 48 h (Mean difference at 0 h = −20.28 U/L and 48 h = −20.01 U/L) with a higher transient decline at 24 h timepoint (−54.08 U/L). Similarly, mean difference in LDH concentrations showed a decline from 0 h to 48 h in the AshwaSR group (mean difference at 0 h = 14.06 U/L and 48 h = 1.57 U/L), whereas an increase was reported in the placebo group during the same period (mean difference at 0 h = 4.40 U/L and 48 h = 6.40 U/L). Notably, at the 24 h post-exercise timepoint for LDH concentrations, an intermediate increase in mean difference in both AshwaSR (22.20 U/L) and placebo (12.38 U/L) groups, followed by a subsequent decline at 48 h was observed. All the changes observed above were statistically nonsignificant (*p* > 0.05).

#### 3.1.5 Quality of life by SF-12 health survey

Quality of life assessed using SF-12 health survey revealed that the participants from AshwaSR group reported statistically significantly increased mean scores for mental and physical components at day 91 compared to baseline (*p* < 0.001) while those from placebo group reported statistically significantly decreased mean scores for both components at day 91 compared to baseline (*p* < 0.001). A between-group comparison demonstrated that the mean changes in scores from baseline for both mental and physical components in the test group were statistically significantly higher than that observed in the placebo group (*p* < 0.001) (Figure 5)

**Figure 5:**
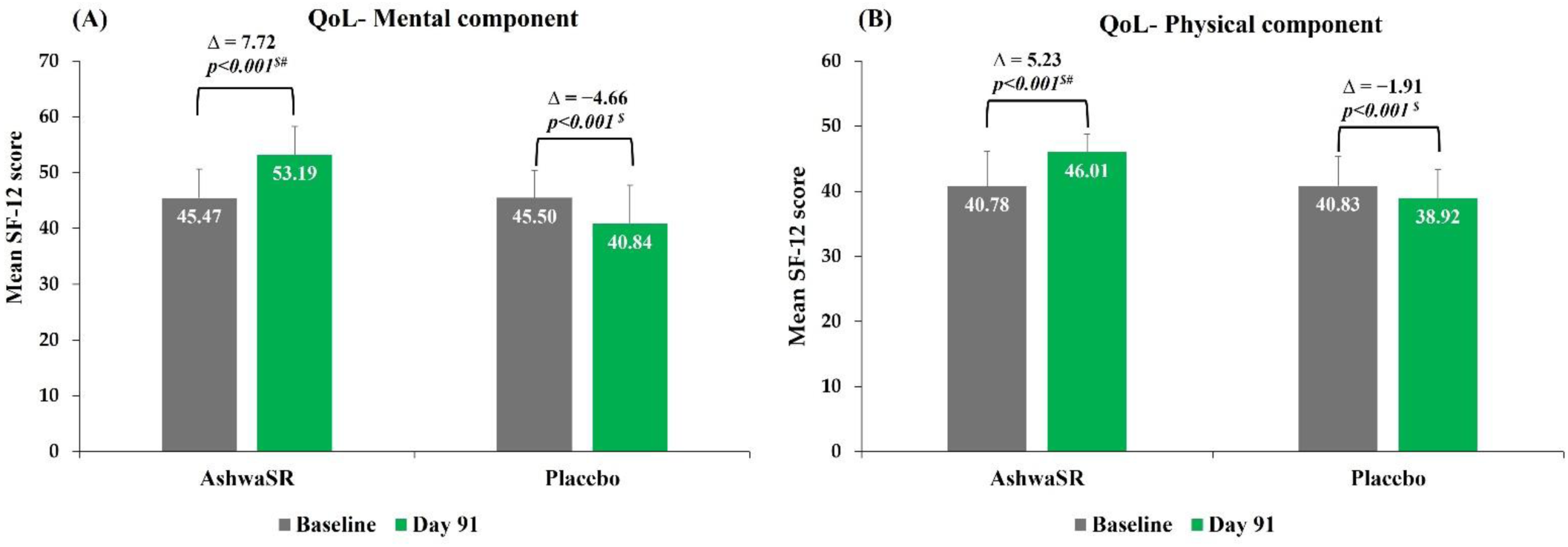
Comparison of mean change in SF-12 scores from baseline to day 91 between AshwaSR and placebo. (A) QoL – Mental component; (B) QoL – Physical component. Data are presented as mean and SD (Error bars represent SD). Δ represents mean change from baseline to the follow-up timepoint (days 91). *p*-value < 0.001 indicates statistically significant difference. ^#^ shows comparison of mean scores between baseline and day 91 (*p*-value derived from Paired t-test). ^$^ shows comparison of the mean change from baseline to day 91 between AshwaSR and placebo (*p*-value derived from ANCOVA, with baseline as covariate). Abbreviations: ANCOVA, analysis of covariance; QoL, quality of life; SD, standard deviation.

### 3.2 Subject’s and physician’s Global assessment of therapy

Subject’s and physician’s global assessment of therapy indicated greater overall acceptability of AshwaSR from both participants’ and physicians’ perspectives. In the AshwaSR group, a majority of participants (n = 37, 90.24%) rated the treatment as Very good/Good. This favorable perception was consistent with physician’s assessment, where physician rated that for majority of participants (n = 40, 97.56%) AshwaSR treatment was Very good/ Good. However, in placebo group, a contrasting trend was observed in these ratings.

### 3.3 Safety assessment

A total of 16 participants reported transient, self-limiting adverse events during the study. Of these, six adverse events were reported by participants from Ashwa SR group (gastritis [n = 03]; Myalgia [n = 03]) while ten were reported by participants from placebo group (gastritis [n = 03]; Myalgia [n = 01]; Backache [n = 02]; Knee pain [n = 02]; and Headache [n = 02]). All adverse events were mild in intensity, resolved without sequelae, and were deemed unlikely to be related to the investigational product based on causality assessment. Clinical safety assessment showed no clinically significant changes from baseline to the end of the study in systolic or diastolic blood pressure, pulse rate, body temperature, oxygen saturation, respiratory rate, or findings from physical examinations in either of the study groups. Similarly, no clinically meaningful abnormalities were observed in laboratory parameters throughout the study period. Overall, both interventions were well tolerated.

## 4 Discussion

Resistance exercise induces mechanical strain on skeletal muscle, leading to activation of intracellular signaling pathways that promote protein synthesis and eventually, muscle building and strengthening. However, during the initial phases of resistance training, particularly in untrained individuals, unfamiliar mechanical loading can induce muscle soreness, fatigue, and delayed recovery. These effects are largely attributed to exercise-induced muscle damage and associated nociceptive responses. When the imposed training load exceeds the body’s adaptive capacity, it may result in physiological stress ultimately leading to reduced adherence to exercise programs (30, 31). Therefore, nutraceuticals that facilitate rapid recovery from this exercise-induced muscle damage while improving muscle strength are of considerable importance, particularly among individuals adopting structured fitness regimens for long-term health and longevity.

In this trial, we evaluated the efficacy and safety of a sustained-release ashwagandha root-only extract (AshwaSR 300 mg), administered once daily for 90 days, in recreationally active healthy individuals. The key observations demonstrated that compared with placebo, the AshwaSR supplementation was efficacious in significantly improving muscle strength and cardiorespiratory endurance, with effects evident from day 30 and sustained through the end of the study. Additionally, favorable changes in testosterone concentration and muscle damage recovery-related biomarkers were observed at the end of the study. These objective outcomes were further supported by subjective assessment wherein participants from AshwaSR group reported significantly improved QoL than placebo group. Collectively, these findings suggest that AshwaSR may serve as an effective ergogenic nutraceutical adjunct to exercise training by enhancing training adaptation, facilitating recovery from muscle damage, and supporting anabolic responsiveness.

Resistance training alone is known to improve muscle strength, which is also observed in the present trial, where participants from test as well as placebo groups showed significantly enhanced muscle strength from baseline. However, AshwaSR supplementation resulted in significantly greater gains in 1-RM bench press performance compared with placebo suggesting two important facts; i) an additional ergogenic benefit of ashwagandha root extract beyond exercise training alone and ii) its ability to enhance upper body strength adaptation during resistance training. Notably, the magnitude of improvement was >2-fold higher than that observed in placebo, starting from as early as day 30 and maintained till end of the study. These beneficial effects may be attributed to the adaptogenic properties of ashwagandha, which substantially enhanced muscle strength by making the resistance training more tolerable through improved recovery and aiding better adaptability to the stimulus of mechanical load on the skeletal muscle. AshwaSR may facilitate a more favorable biochemical environment for muscle adaptation and recovery through indirect mechanisms, including modulation of the hypothalamic–pituitary–adrenal (HPA) axis, improvement of quality of rest required for recovery, and reduction in perceived exertion and pain sensitivity (21, 22, 32, 33). Collectively, these effects may support better recovery between training sessions, improve adherence to exercise regimens, and ultimately contribute to greater gains in muscle strength over time.

Similar trends of improvement in muscle strength were reported in physically active adults receiving conventional standardized ashwagandha extracts (13, 14, 34). For instance, a study in recreationally active men demonstrated that supplementation with an aqueous extract of ashwagandha at a dose of 500 mg/day for 12 weeks significantly improved 1-RM bench press strength (14). Similarly, a randomized controlled trial reported that administration of a standardized ashwagandha extract at 300 mg twice daily for 8 weeks resulted in significantly greater increases in upper body strength compared with placebo (46.05 kg vs. 26.42 kg; *p* = 0.001) in healthy young men undergoing resistance training (13). However, not all studies have reported consistent findings. A randomized clinical trial in physically active adults reported non-significant differences in strength gains between ashwagandha (600 mg/day for 8 weeks) and placebo groups (mean change: 11.0 kg vs. 8.0 kg; *p* = 0.176), suggesting that efficacy may vary depending on population characteristics, study design, or formulation (35). In this regard, an important distinction of the present trial is the use of a sustained-release formulation at a low dose with only once-a-day administration. Most prior studies have employed immediate-release extracts administered once or twice daily. In contrast, AshwaSR is designed to provide gradual release of active constituents over an extended period, resulting in more stable physiological exposure for longer duration. The sustained-release profile of actives in the AshwaSR formulation has been demonstrated in a published pharmacokinetic study, in which the test formulation showed greater withanolides bioavailability and a longer elimination half-life following once-daily administration compared with the reference product (a marketed ashwagandha root extract capsule) (20). This delivery profile may be advantageous during long-term exercise programs, where sustained support for recovery and adaptation is desirable.

Cardiorespiratory endurance, a well-known component of physical fitness, was another important outcome evaluated in the present trial. Resistance training has also been shown to improve VO_2_max through adaptations such as enhanced cardiac-muscle function, increased blood volume, and improved oxygen-carrying capacity (36, 37). In this trial, AshwaSR supplementation resulted in a significant improvement in cardiorespiratory endurance, with benefits evident from day 30 onward and maintained throughout the study period. Notably, the magnitude of improvement in VO₂max in the AshwaSR group was approximately 1.5-fold greater than that observed in the placebo group.

These findings suggest that AshwaSR may enhance tolerance to aerobic exertion, potentially by facilitating recovery and improving physiological adaptability to progressively increasing cardiorespiratory demands. These observations align with published literature wherein ashwagandha supplementation was shown to improve cardiorespiratory endurance in athletic as well as non-athletic population (13, 16, 35, 38–41). A randomized clinical trial involving physically active adults reported that supplementation with ashwagandha extract at a dose of 300 mg twice daily for 8 weeks resulted in an approximately 8% improvement in cardiorespiratory endurance from baseline compared with placebo, although between-group differences were not significant at the 4-week follow-up (35). However, the present trial demonstrated a substantially greater and earlier improvement over the 90 days study duration, with an approximately 47% increase in VO₂max observed as early as day 30 following once-daily administration of AshwaSR 300 mg, compared to that observed in placebo group. Interestingly, this magnitude of improvement exceeds that reported in the aforementioned study at 8 weeks and appears notable given the lower dosing frequency used in the present trial. Moreover, previous studies have generally reported more modest improvements in VO₂max, typically ranging between 8% and 16% after 8 weeks of supplementation (35, 39, 40).

Taken together, these findings suggest that the AshwaSR, may offer advantages over conventional ashwagandha preparations by producing earlier, larger, and potentially more sustained improvements in cardiorespiratory endurance. These observations also highlight that the efficacy of ashwagandha supplementation may be influenced by formulation characteristics, including delivery technology, purity and quality of the extract. The simultaneous improvement in muscular strength and aerobic capacity observed in the present trial are particularly relevant for recreationally active individuals, as these components collectively contribute to overall fitness, exercise performance, and long-term adherence to exercise.

Testosterone is an important anabolic hormone involved in muscle protein synthesis, and recovery; and maintenance of an optimal testosterone balance is vital for muscle health, metabolic function, and overall quality of life (42). In the present trial, male participants receiving AshwaSR reported significant increases from baseline in total, and free testosterone concentrations. Although between-group differences did not reach statistical significance and testosterone concentrations remained within normal physiological ranges in both groups, the magnitude of change was numerically greater in the AshwaSR group than placebo, suggesting a favorable effect on anabolic adaptation. These findings add corroborative evidence to existing literature wherein significantly greater increases in testosterone concentration were reported among resistance-trained men supplemented with ashwagandha extract compared to placebo (13). In the present trial, female participants from the test group reported non-significant changes at the end of the intervention period whereas in the placebo group, significantly high testosterone levels were reported at the end of the study which crossed upper limit of reference range in case of free testosterone. These findings indicate that test product exerted its adaptogenic effect in females by preventing excessive androgen (total and free testosterone) elevation during intensive training and retained their concentrations within normal range. This also suggests a gender-specific adaptogenic property of AshwaSR, supporting performance without hormonal dysregulation in females. However, these findings are preliminary and may be validated through further clinical studies.

In the present trial, evaluation of muscle damage biomarkers at post-exercise time points was conducted to assess the effect of AshwaSR on recovery dynamics and adaptation to training. Participants receiving AshwaSR demonstrated lower CK and LDH levels at 48 h post-exercise at the end of the study compared with the placebo group, suggesting improved recovery from exercise-induced muscle damage and better physiological adaptability of participants towards workout in AshwaSR group. Importantly, AshwaSR did not appear to blunt the physiological stress associated with resistance exercise but rather promoted a favorable recovery profile characterized by lower late-phase reductions of muscle damage markers. Although transient increases in LDH concentrations are expected following resistance exercise, attenuation of excessive elevations or faster normalization may reflect improved recovery capacity (43). Previous evidence in untrained individuals has shown that the maximum serum concentrations of CK were noted at 48 hours after exercise and maximum serum concentration of LDH was dependent on rest interval between sets and load during the exercise, with lower load resulting in peak at 72 hours and higher load resulting in peak at 24 hours (44). The findings of the present study are consistent with these observations and support the potential role of AshwaSR in enhancing recovery and adaptability to structured exercise training.

Beyond physiological outcomes, subjective evaluation of participants’ wellbeing in this study demonstrated that AshwaSR group reported significantly better improvement in both the physical and mental domains of SF-12 health survey questionnaire than placebo. Particularly, by day 90, participants in the placebo group reported contrasting trend compared to test group, with significant decrease observed in SF-12 scores. This divergence offers compelling evidence that AshwaSR effectively enhances both mental and physical health-related QoL, aligning with the adaptogenic and performance-supporting properties of Ashwagandha. These improvements may be attributed to the withanolide-rich phytochemical profile of AshwaSR, which has been associated with GABAergic and serotonergic modulatory activity, along with antioxidant and anti-inflammatory effects that collectively support emotional stability, psychological resilience, and physical vitality (45). These effects may indirectly enhance training consistency and perceived capacity to exercise. In addition, subjects and physicians’ global assessment of therapy reflected a better acceptability of AshwaSR for its benefits from both subject’s and physician’s perspective. In safety assessment, AshwaSR was well tolerated throughout the study. Reported AEs were mild, transient, and considered unrelated to the investigational products. These findings are consistent with the generally favorable safety profile reported for standardized ashwagandha root extracts in previous human trials (35, 46).

To our knowledge, this is the first trial to evaluate the effect of a sustained-release ashwagandha root extract supplementation on both muscle strength and cardiorespiratory endurance, over a relatively longer period of 3 months, in recreationally active healthy male and female adult participants. The study duration also represents one of the longest interventions administered to evaluate the role of a sustained-release ashwagandha formulation in sports nutrition. Furthermore, implementation of a standardized and closely supervised exercise regimen by trained study personnel ensured consistency in training exposure across participants and minimized variability arising from differences in workout practices. Notably, the test formulation demonstrated a gender-specific adaptogenic profile with testosterone concentrations in women maintaining within normal physiological ranges despite improvements in exercise performance, thereby reinforcing its suitability and safety for both male and female recreationally active adults. Lastly, the multidimensional evaluation encompassing physical performance, endocrine parameters, muscle damage biomarkers, and participant-reported outcomes provides a comprehensive assessment of the efficacy of the test product and addresses an underexplored area of nutraceuticals in sports performance research.

Few limitations acknowledged include the study population limited to recreationally active healthy adults, and exploring effects of AshwaSR in various athletic as populations, well as non-athletic such as sedentary populations, and older adults, may help further establish the benefits of AshwaSR across diverse exercise settings. Since AshwaSR is available in both 150 mg and 300 mg strengths, evaluation of multiple dose levels could provide further insight into the dose-response relationship and sustainability of efficacy on these study endpoints. Additionally, mechanistic biomarkers related to inflammation, oxidative stress, and cortisol regulation or body composition imaging may be evaluated in future studies.

## 5 Conclusion

In conclusion, supplementation with sustained-release ashwagandha root extract (AshwaSR) 300 mg once daily for 90 days was safe and effective in improving muscular strength, cardiorespiratory endurance, and post-exercise muscle recovery, gen-der-specific regulation of testosterone and mental and physical wellbeing leading to overall improvements in physical performance in recreationally active healthy adults. These findings point towards the evidence favoring sustained-release profile of AshwaSR as a suitable alternative to support exercise adaptation and serve as a potential ergogenic nutraceutical aid for the individuals engaged in fitness training.

## 7 Author Contributions

Conceptualization, S.T., R.S., and A.L.; formal analysis and data curation, G.H., P.D, and S.S; investigation, G.H. and S.S.; writing—original draft preparation, S.T. and A.L. ; writing—review and editing, all authors; visualization, R.S. and S.T.; supervision, S.T., A.L. and P.D.; project administration, A.L. and S.S.; funding acquisition, R.S. All authors have read and agreed to the published version of the manuscript.

## 9 Acknowledgments

The authors would like to thank all the study participants who were enrolled in this study. The medical writing and editorial support were provided by Tejal Dhotre of Nutriventia Private Limited.

## 10 Data availability statement

The data that support the findings of this study are available from the corresponding author upon reasonable request.

## 11 Supplementary Material

Not applicable.

